# K-means clustering of hyperpolarised ^13^C-MRI identifies intratumoural perfusion/metabolism mismatch in renal cell carcinoma as best predictor of highest grade

**DOI:** 10.1101/2024.05.06.24306829

**Authors:** Ines Horvat-Menih, Alixander S Khan, Mary A McLean, Joao Duarte, Eva Serrao, Stephan Ursprung, Joshua D Kaggie, Andrew B Gill, Andrew N Priest, Mireia Crispin-Ortuzar, Anne Y Warren, Sarah J Welsh, Thomas J Mitchell, Grant D Stewart, Ferdia A Gallagher

**Affiliations:** Department of Radiology, University of Cambridge, UK; (I.H.M); (A.S.K.); (M.A.M); (J.D.); (E.S.); (J.D.K.); (A.B.G.); (A.N.P); (F.A.G.); Department of Radiology, Royal Papworth Hospitals NHS Foundation Trust, Cambridge, UK; Department of Radiology, Addenbrooke’s Hospital, Cambridge University Hospitals NHS Foundation Trust, UK; Department of Oncology, University of Cambridge, UK; (M.C.O.); Department of Pathology, Addenbrooke’s Hospital, Cambridge University Hospitals NHS Foundation Trust, UK; (A.Y.W.); Pinto Medical Consultancy, UK; (S.J.W.); Department of Surgery, University of Cambridge, UK; (T.J.M); (G.D.S.)

## Abstract

**Purpose:** Conventional renal mass biopsy approaches are inaccurate, potentially leading to undergrading. This study explored using hyperpolarised [1-^13^C]pyruvate MRI (HP ^13^C-MRI) to identify the most aggressive areas within the tumour of patients with clear cell renal cell carcinoma (ccRCC).

**Experimental design:** Six patients with ccRCC underwent presurgical HP ^13^C-MRI and conventional contrast-enhanced MRI. Three k-means clusters were computed by combining the *k*_PL_ as a marker of metabolic activity, and the ^13^C-pyruvate signal-to-noise ratio (SNR_Pyr_) as a perfusion surrogate. Combined clusters were compared to those derived from individual parameters and to those derived from percentage enhancement on nephrographic phase (%NG). The diagnostic performance of each cluster was assessed based on its ability to predict the highest histological tumour grade in postsurgical tissue samples. Tissues were further subject to MCT1 staining, RNA and whole-exome sequencing.

**Results:** Forty-four samples were collected in total. The clustering approach combining SNR_Pyr_ and *k*_PL_ demonstrated the best performance for predicting highest tumour grade: specificity 85%; sensitivity 64%; positive predictive value 82%; and negative predictive value 68%. Epithelial MCT1 was identified as the major determinant of the HP ^13^C-MRI signal. The perfusion/metabolism mismatch cluster showed increased expression of metabolic genes and markers of aggressiveness, which may be due to genetic divergence.

**Conclusions:** This study demonstrates the potential of using HP ^13^C-MRI-derived metabolic clusters to identify intratumoral variations in tumour grade with high specificity. This work supports the use of metabolic imaging to guide biopsies to the most aggressive tumour regions, which could potentially reduce sampling error.

## Introduction

Renal cell carcinoma (RCC) represents 2% of cancer diagnoses globally, with an increasing incidence which is partly attributed to incidental detection on imaging^1^. However mortality has not decreased over the last 50 years, despite this earlier detection and improved treatment, with RCC remaining the most lethal urological malignancy^2^. A major diagnostic challenge is differentiating between aggressive small renal masses and indolent lesions, as well as accurately determining tumour grade before surgery^3^. Although renal mass biopsy is a key tool for discriminating radiologically indeterminate renal lesions, it is unrepresentative of the whole tumour and may be subject to sampling error, which can result in undergrading of heterogeneous lesions, potentially delaying surgery^4–6^. Therefore, there is a pressing need for novel non-invasive methods to characterise renal masses more accurately and to stratify tumours.

Metabolism may be particularly useful for phenotyping RCCs which are characterised by a high degree of metabolic reprogramming driving tumour formation^7^. For example, in the most common and aggressive RCC subtype, clear cell RCC (ccRCC), the inactivation of the von Hippel-Lindau (*VHL*) gene leads to stabilisation of hypoxia-inducible factor (HIF) with downstream activation of angiogenesis, glycolysis, and suppression of the oxidative metabolic pathways^8^. Intratumoral metabolic heterogeneity within RCC has been observed across grades and within spatially separated tumour regions on *ex vivo* molecular analyses^9,10^. Assessment of this heterogeneity with *in vivo* imaging such as [^18^F]fluorodeoxyglucose (FDG)-positron emission tomography (PET) has been limited by the renal excretion of the FDG-tracer^11^. Hyperpolarised [1-^13^C]pyruvate MRI (HP ^13^C-MRI) is an emerging clinical imaging technique to non-invasively probe tumour lactate formation from injected pyruvate, which offers the potential to stratify ccRCC based on metabolism using a non-radioactive endogenous metabolite^12^. For instance, recent studies have shown the potential of the technique in stratifying tumours based on higher WHO/ISUP grade^13,14^, with increased pyruvate metabolism correlating with the pyruvate importer monocarboxylate transporter 1 (MCT1) as an independent marker of poor prognosis in ccRCC^14,15^.

In this study we have explored the role of metabolic imaging in assessing tumour aggressiveness by combining metrics of perfusion and metabolism, acquired using HP ^13^C-MRI, to assess whether a combined approach may be more powerful in assessing grade than the individual metrics alone, or than the conventional contrast-enhanced proton (^1^H) MRI. K-means clustering was used as an unsupervised learning algorithm that clusters pixels based on intensity and spatial positioning, to disaggregate areas of metabolic similarity in the tumour, as has been used in ^1^H-MRI to evaluate distinct tumour regions^16^. The results have shown the potential of metabolic MRI to detect areas of increased tumour aggressiveness, which could be used to guide a biopsy to the most aggressive areas within an individual tumour in the future.

## Patients & methods

### Ethics and Recruitment

Patients were prospectively recruited and provided written informed consent to the following ethically approved studies: MISSION-Ovary Substudy Renal (ClinicalTrials.gov Identifier NCT03526809), and ARTIST (NCT04060537). This cohort of patients overlaps with previously published work^14^. Key inclusion criteria were: age ≥ 18 years old, known renal mass scheduled for partial or radical nephrectomy, confirmed ccRCC at histopathology, and successful HP ^13^C-MRI acquisition. Exclusion criteria were uncontrolled metabolic disorders, contraindications to gadolinium (Gd)-containing contrast agents, and inability to undergo MRI. Seventeen patients with a renal mass suspicious for ccRCC were prospectively enrolled and underwent presurgical imaging; of these, thirteen had ccRCC on final histology and six successfully underwent HP ^13^C-MRI acquisition.

### MRI acquisition and processing

The ^13^C-pyruvate injection and the HP ^13^C-MRI acquisition were performed as described in previously published work, using the 3T MRI (MR750, GE Healthcare, Waukesha, WI, USA) and a ^13^C-tuned clamshell for transmission (GE Healthcare, Waukesha, WI, USA) and an 8-channel array coil for reception (Rapid Biomedical, Rimpar, Germany)^14,17^. HP ^13^C-MRI data were reconstructed using in-house MATLAB (Mathworks, Massachusetts, USA) scripts where metabolite spectra from individual timepoints were summed together, and the complex imaging data from the 8-channels of the abdominal coil were combined using the singular value decomposition approach^18^. The *k*_PL_ was calculated using a two-site exchange kinetic model in the frequency domain^19^. Signal-to-noise ratio of ^13^C-pyruvate (SNR_Pyr_) for each ROI was calculated based on Equation 1, with mean and standard deviation (SD) of the signal intensity (SI) of noise measured outside the patient.

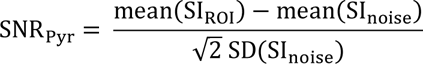

Equation 1: SNR_Pyr_ calculation from individual ROIs. SI_ROI_ = signal intensity in the ROI, SI_noise_ = signal intensity in the noise ROI.

^1^H-MRI sequences, including T_1_- and T_2_-weighted (T_1_w, T_2_w) and Gd-contrast enhanced imaging were acquired after replacing the ^13^C-tuned coil with the 32-channel cardiac ^1^H-array coil (GE Healthcare, Wisconsin, USA), and processed as described previously^14^. Percentage nephrographic enhancement (%NG) as a measure of vascular permeability was calculated by subtracting the non-enhanced T_1_-weighted sequence from the nephrographic contrast-enhanced phase, the latter defined as the timepoint of maximal and homogeneous enhancement across renal parenchyma, corresponding to ∼100 s after the start of Gd-injection^20^. To match the imaging planes and correct for any potential movement, the final ^1^H-maps were reoriented and manually registered to match the HP ^13^C-MRI using ITK-SNAP 4.0^21^ and in-house developed MATLAB scripts. Using the OsiriX Lite 12.0.3 (Pixmeo SARL, Switzerland), the regions of interest (ROIs) of whole tumours were drawn on axial T_1_w LavaFlex images by avoiding cystic/necrotic areas via simultaneous inspection of T_2_w and Gd-enhanced images.

### K-means clustering

ROIs drawn on anatomical T_1_w images were applied to HP ^13^C-MRI and %NG images to create masked maps as shown in Figure 1. Masked tumour regions were extracted, and clustering was performed using MATLAB using both combined HP ^13^C-MRI inputs [SNR_Pyr_ + *k*_PL_] and individual inputs of the HP ^13^C-MRI data and %NG. Three clusters were created for each clustering approach, whereas voxels with poor signal-to-noise ratio indicated a necrotic area and were matched to background. A sorting algorithm was used following clustering to arrange the habitats in order of magnitude to ensure that the highest cluster always corresponded to the highest magnitude of imaging data.

**Figure 1:**
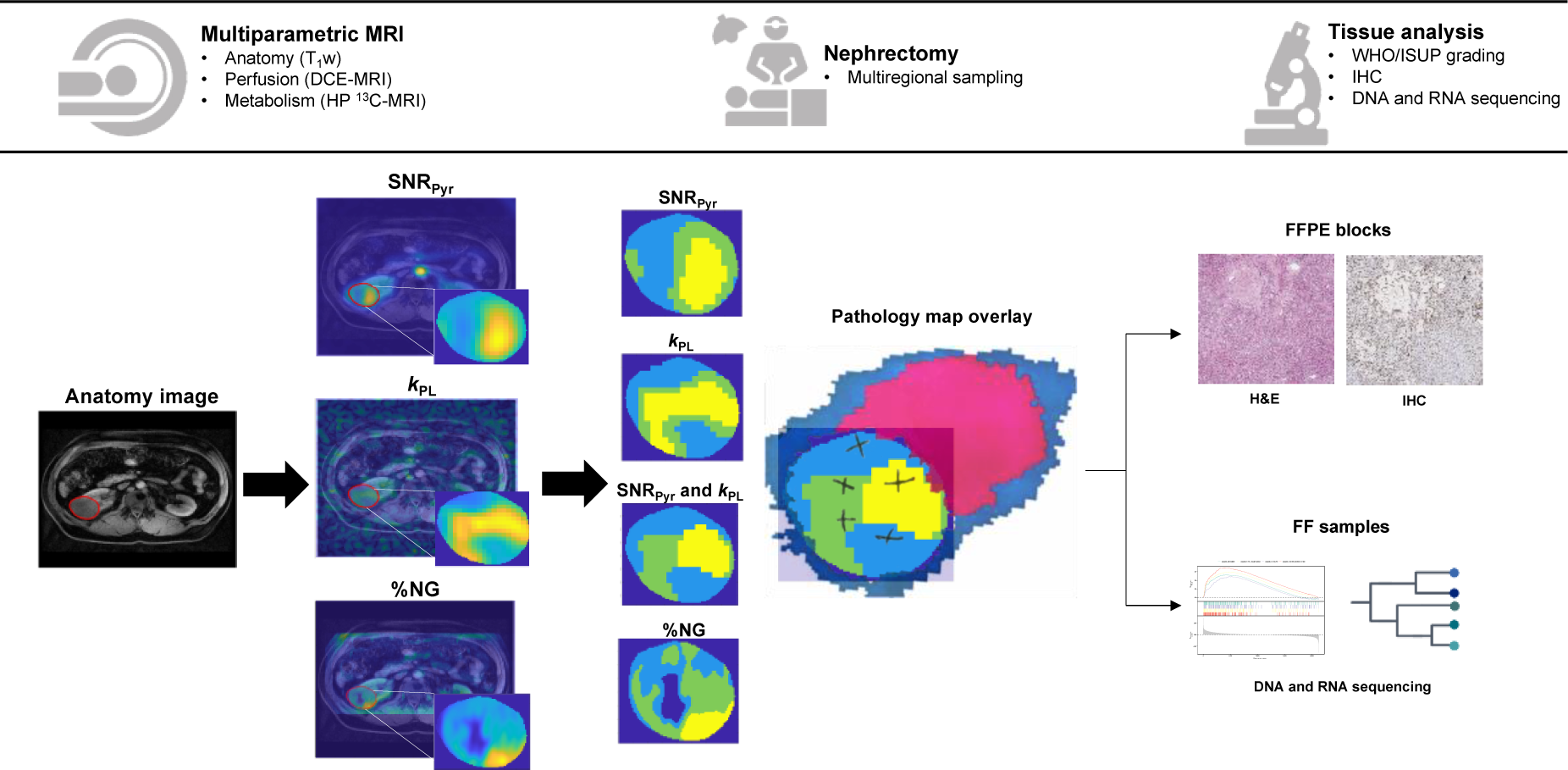
Workflow of the study. Patients underwent multiparametric MRI including anatomical imaging (T_1_w), Gd-enhanced MRI, and HP _13_C-MRI. Region of interest (ROI) was delineated on anatomy image, shown as red circle surrounding tumour. SNR_Pyr_ and *k*_PL_ maps from HP _13_C-MRI, and the %NG map calculated from Gd-enhanced MRI, were co-registered to anatomy and are shown as overlays on top of T_1_w image. ROIs were extracted from each map and k-means clustering was applied to define three clusters for all individual parameters, and the combination of normalised [SNR_Pyr_ + *k*_PL_]. The colour code for the three clusters is as follows: light blue represents the lowest, green the medium, and yellow the highest mean value, while dark blue denotes the background or any voxels with poor signal-to-noise ratio which was matched to background. After nephrectomy each tumour was sliced at a specified location to match the imaging acquisition plane using the tumour mould, and the location of the biopsies was recorded on the patient-specific pathology maps. These were overlaid on top of each clustering result, with the normal kidney in purple, perirenal fat in blue, and the location of the biopsies drawn as crosses. Each biopsy was split in two, with one half undergoing FFPE preparation, with the other half being fresh frozen (FF). Sections from FFPE were stained with H&E for WHO/ISUP grading, IHC for MCT1, while FF samples underwent DNA and RNA sequencing.

### Tissue sampling and analyses

To enable co-registration of the biopsies to the MRI images, the patient-specific pathology sampling maps were produced from MRI biomarker maps and sliced with the assistance of a 3D printed tumour mould as described previously^22^. Four to fourteen multiregional samples were collected from viable tumour regions by avoiding visible cystic/necrotic areas, and each biopsy was split into half with one part being formalin-fixed paraffin-embedded (FFPE) and the other half flash-frozen. A consultant uropathologist determined the WHO/ISUP grade on H&E of all the biopsies with >75% tumour cellularity. Immunohistochemical (IHC) staining for the pyruvate transporter (monocarboxylic acid transporter 1, MCT1) was performed on the FFPE sections and analysed as described previously^14^, while fresh frozen samples underwent RNA sequencing and whole-exome sequencing (WES), as described below.

### RNA sequencing and differential expression analysis

Extracted RNA was isolated and prepared using previously described workflows^23^. Sequencing was performed on the HiSeq4000 platform (Illumina, Inc., San Diego, California, USA). Mapping and quality control used the RNA STAR mapping algorithm^24^. HTSeq 0.7.2 was used to generate RNA count data^25^, and differential expression analysis across imaging clusters was performed using the DESeq2 package 1.34.0^26^ and Genekitr package 1.2.2^27^ gene set enrichment analysis (GSEA), enriched for curated The Molecular Signatures Database (MSigDB) KEGG gene sets^28,29^. Model matrix was constructed to correct for patient batch effects due to differential numbers of biopsies per tumour.

### Cell signal analysis and deconvolution from bulk RNA sequencing

To estimate the composition of tumours and infer the most conserved cellular signals observed in the bulk RNA sequencing (RNAseq) data, an adult kidney reference derived from normal kidneys of patients with RCC was used^28^. BayesPrism^29^ was used to predict the cellular composition of the tumours from bulk RNA using single cell references. To predict the cellular composition of tumours we collated the reference cell subtypes according to their major lineage e.g. leukocytes, myeloid, endothelial, epithelial, fibroblast, rather than using the finely annotated subtypes. The collation to major lineage types helped alleviate batch effects and the difficulty in discerning the presence of subtly different cell subtypes from bulk RNA data.

### Mutation calling from whole-exome sequencing

DNA sequencing was performed on the Illumina HiSeq 4000 platform prior to alignment to the GRCh 37d5 reference genome using the Burrows-Wheeler transform (BWA-MEM)^30^. Single base somatic substitutions nomenclature was based on CaVEMan 1.14.1 (Cancer Variants through Expectation Maximization)^31^. Small insertions and deletions (indels) nomenclature was based on Pindel 3.3.0^32^. Copy-number data were derived from exonic reads using the ASCAT v2.3 algorithm^33^.

### DNA mutational clustering

Mutation counts at each loci were calculated across all samples using AlleleCounter^34^. Mutations were clustered using a Bayesian Dirichlet based algorithm as described previously^35^. Briefly, the expected number of reads for a given mutation present in one allelic copy of 100% of tumour cells may be estimated based upon the ASCAT derived tumour cell fraction, the copy number at that locus, and the total read depth. The fraction of cells carrying a given mutation is then modelled by a Dirichlet process with an adjustment for the decreased sensitivity in identifying mutations in lower tumour fractions. Mutations were thus assigned to clusters according to the calculated fraction of clonality. The hierarchical ordering of these clusters was determined by applying the pigeonhole principle to generate consensus phylogenies.

### Statistical analysis

Statistical analysis was performed in GraphPad (Prism 10, San Diego, USA). All biopsies were assigned to a cluster based on the co-registration of the MRI and the pathology. The diagnostic performance for predicting the highest-grade within the tumour was calculated for all clusters derived from individual and combined clustering. Diagnostic performance was assessed using sensitivity, specificity, and positive and negative predictive values (PPV, NPV), and the area under curve (AUC) of the receiver operating characteristic (ROC) curves served as a consistency measure. MCT1 and cell-type compositions were compared across clusters by applying a one-way ANOVA test with Holm-Šidak’s multiple comparison correction for normally distributed data, and the Kruskal-Wallis test with Dunn’s multiple comparisons correction for nonparametric data, as tested by Shapiro-Wilk test for normality. Binary comparisons were correspondingly performed using unpaired Student’s t-test and Mann-Whitney U test for gaussian and non-gaussian data, respectively. *P* value < 0.05 was set as a cut-off for statistical significance.

### Data Availability

All data that support the findings of this study are available on reasonable request from the corresponding author, on condition that this will not be used to deanonymize the patients. The data are not publicly available due to them containing information that could compromise research participant privacy and consent.

## Results

### Study workflow and patient clinical characteristics

The study workflow is shown in Fig. 1 and the summary characteristics of the six patients is presented in Table 1. Median age was 59.5 years (range 51-69), one patient was female. The tumour size ranged from 4.0 to 13.3 cm. Metastatic disease was present in one patient. Median time between imaging and surgery was 16.5 (range 1-119) days. In total, forty-four multiregional samples were post-operatively collected, of which two were excluded due to insufficient tumour cellularity.

**Table 1:**
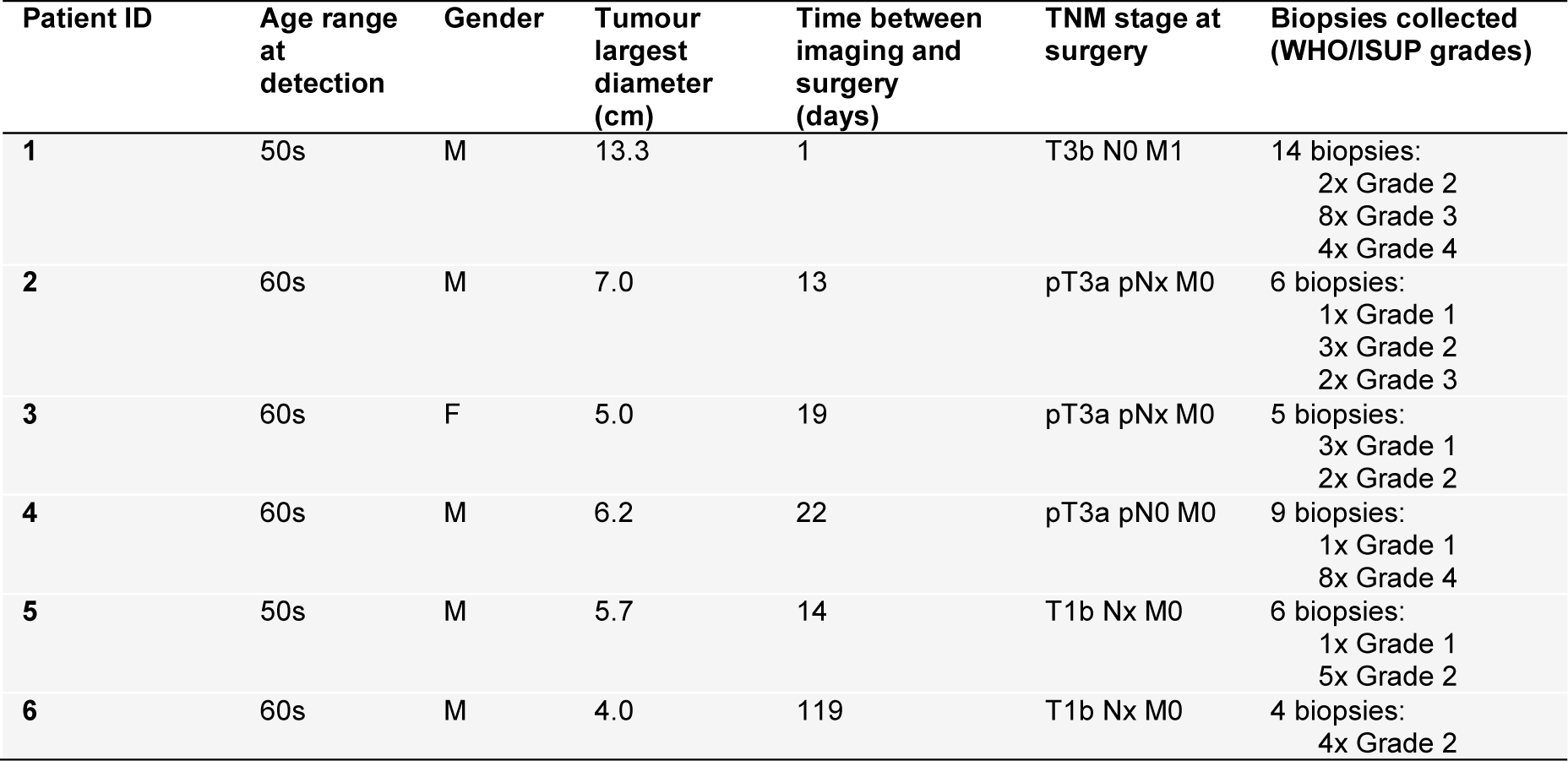
Patient clinical characteristics.

| Patient ID | Age range at detection | Gender | Tumour largest diameter (cm) | Time between imaging and surgery (days) | TNM stage at surgery | Biopsies collected (WHO/ISUP grades) |
| --- | --- | --- | --- | --- | --- | --- |
| 1 | 50s | M | 13.3 | 1 | T3b N0 M1 | 14 biopsies:<br>2x Grade 2<br>8x Grade 3<br>4x Grade 4 |
| 2 | 60s | M | 7.0 | 13 | pT3a pNx M0 | 6 biopsies:<br>1x Grade 1<br>3x Grade 2<br>2x Grade 3 |
| 3 | 60s | F | 5.0 | 19 | pT3a pNx M0 | 5 biopsies:<br>3x Grade 1<br>2x Grade 2 |
| 4 | 60s | M | 6.2 | 22 | pT3a pN0 M0 | 9 biopsies:<br>1x Grade 1<br>8x Grade 4 |
| 5 | 50s | M | 5.7 | 14 | T1b Nx M0 | 6 biopsies:<br>1x Grade 1<br>5x Grade 2 |
| 6 | 60s | M | 4.0 | 119 | T1b Nx M0 | 4 biopsies:<br>4x Grade 2 |

### K-means clustering of HP ^13^C-MRI detects intratumoral heterogeneity of pyruvate delivery and pyruvate-to-lactate conversion

Fig. 2 shows an overview of the clustering results for all patients. Nephrectomy specimens from patients 1 and 5 were sliced at two different axial levels based on the 3D printed mould which was registered to the HP ^13^C-MRI images as described previously^14^. Slices from patient 1 were sufficiently separated to correspond to two different HP ^13^C-MRI slices (1a and 1b), while the slicing levels of the study patient 5 were both constrained within a single slice on HP ^13^C-MRI images, for which reason biopsies from both levels were overlaid onto the same clustering map. Differences between tissue perfusion (as measured by pyruvate delivery with SNR_Pyr_, and Gd-enhancement with %NG) and tumour metabolism (pyruvate-to-lactate conversion using *k*PL) indicated intratumoral heterogeneity in both, which was captured by combining an [SNR_Pyr_ + *k*PL] clustering approach.

**Figure 2:**
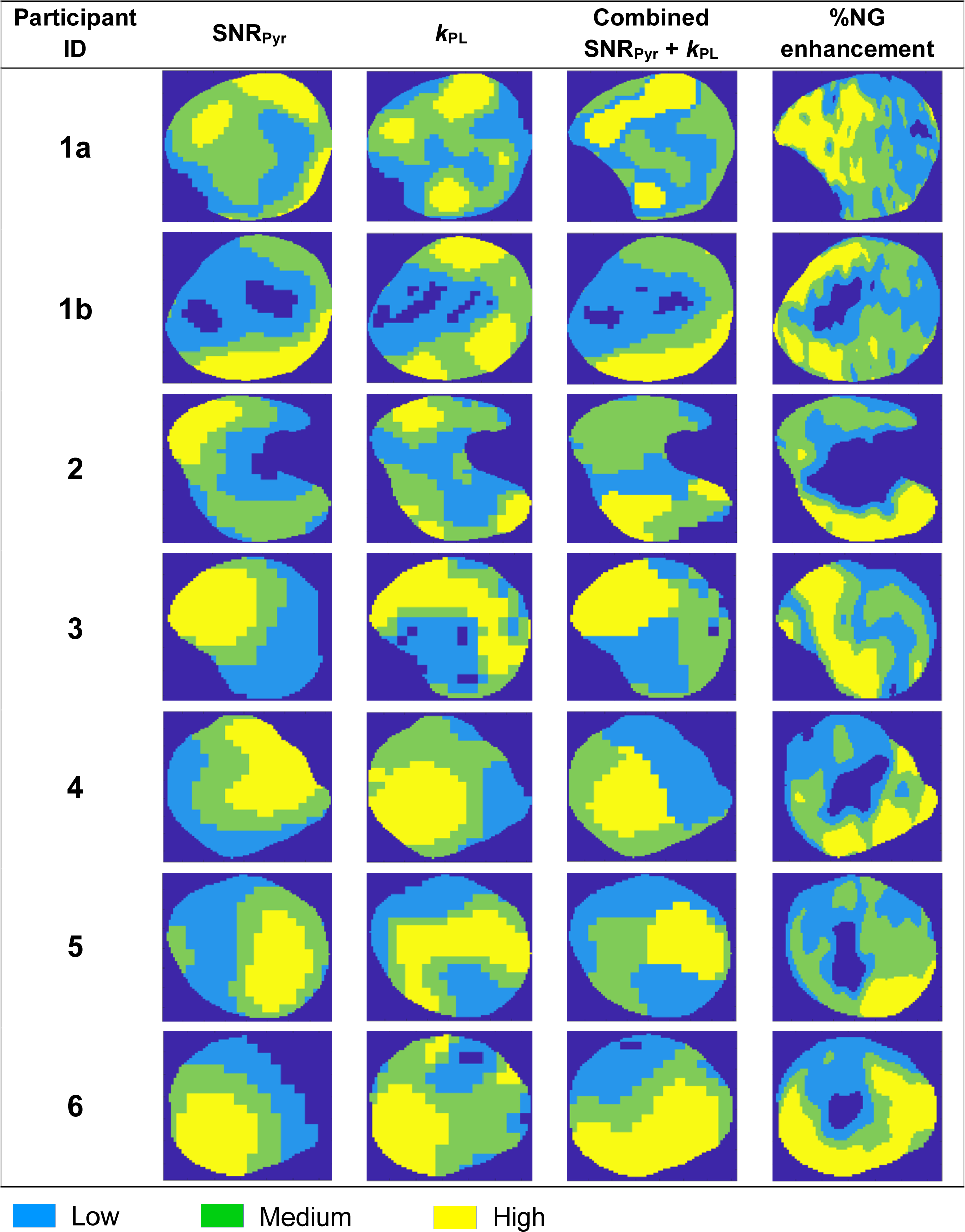
Overview of k-means clustering results in the 6 patients. Tumour from patient 1 was large enough to acquire imaging and biopsies on two levels (1a and 1b), while in other patients only a single slice was imaged and registered to retrieval of post-nephrectomy biopsies. Intratumoral heterogeneity was observed in all clustering approaches.

To quantify the level of heterogeneity of perfusion and metabolism within and between the combined clusters, we compared the normalised individual and combined SNR_Pyr_ and *k*PL means within each combined cluster group (Fig. 3).

**Figure 3:**
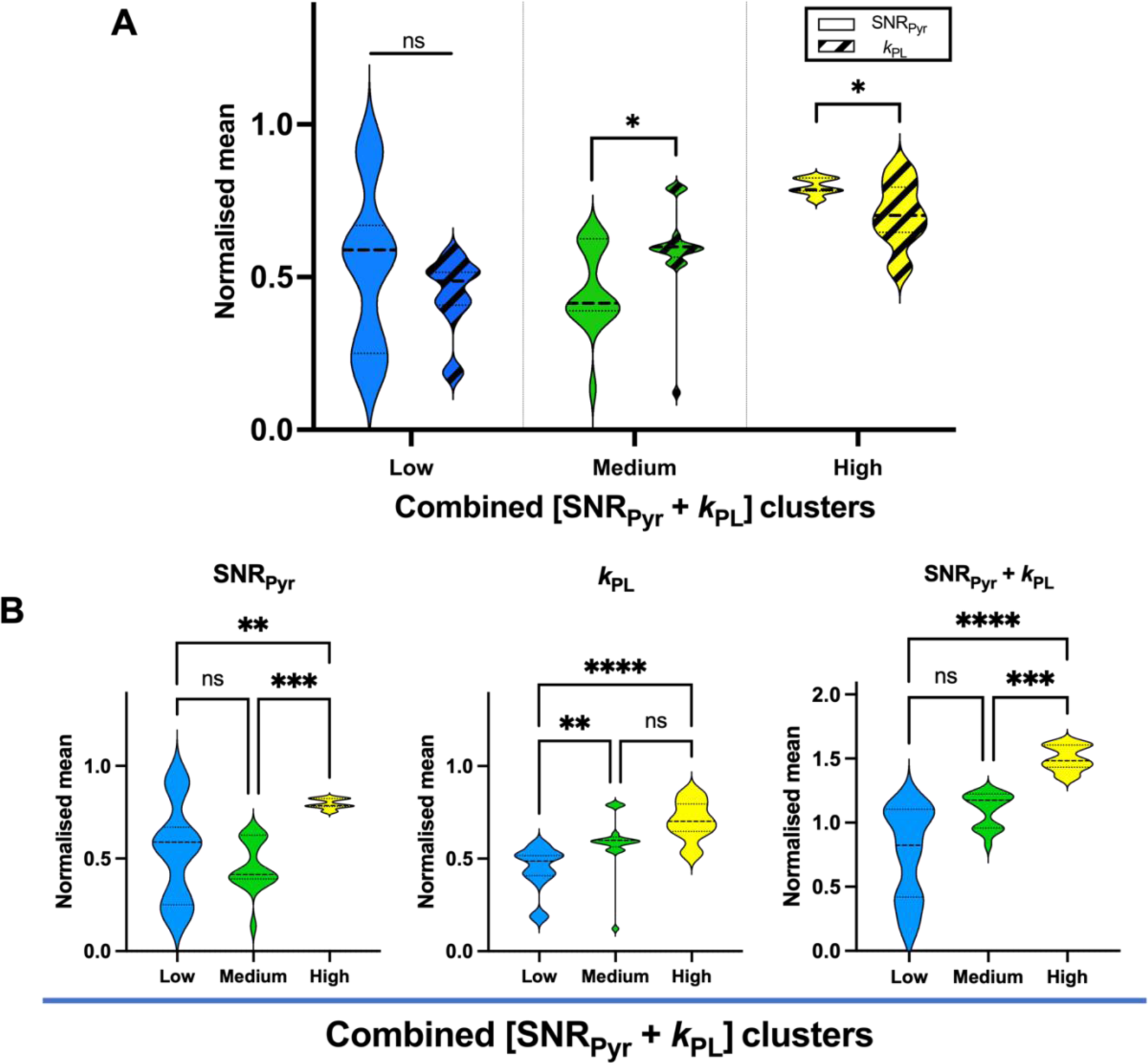
Comparison of imaging parameters within and between combined [SNR_Pyr_ + *k*_PL_] clusters. (A) No difference between SNR_Pyr_ and *k*_PL_ was found in the low combined cluster (Mann-Whitney U *P* = 0.10), while in medium and high combined clusters, the difference between SNR_Pyr_ and *k*_PL_ was significant (Mann-Whitney U *P* < 0.05 in both comparisons): in the medium combined cluster, the SNR_Pyr_ was significantly lower than *k*_PL_ (0.41 vs. 0.60), and in the high combined cluster, the SNR_Pyr_ was significantly higher than *k*_PL_ (0.79 vs. 0.70), suggesting mismatch between pyruvate delivery and conversion to lactate. (B) SNR_Pyr_ across combined clusters was significantly different in the low vs. high (Dunn’s multiple comparisons test adjusted *P* < 0.01), and medium vs. high (*P* < 0.001), but not in low vs. medium clusters (*P* > 0.99). *k*_PL_ across combined clusters was significantly different in low vs. medium (*P* < 0.01), and low vs. high (*P* < 0.0001), but not between medium vs. high clusters (*P* = 0.20). Combined [SNR_Pyr_ + *k*_PL_] was significantly different in low vs. high (*P* < 0.0001), and medium vs. high (*P* < 0.001), but not in low vs. medium clusters (*P* = 0.06).

The medium cluster showed the greatest degree of perfusion/metabolism mismatch with low perfusion and high metabolic conversion, which may represent increased glycolytic metabolism secondary to tumour hypoxia (medium cluster mean *k*_PL_/SNR_Pyr_ ratio = 1.4, compared to 1.0 and 0.9 in the low and high combined clusters, respectively; Fig. 3A). This is analogous to discrepancies found on ^18^F-FDG-PET imaging, which have been reported as a feature of tumour aggressiveness^36–38^.

Fig. 3B shows values of individual SNR_Pyr_, *k*_PL_ and combined [SNR_Pyr_ + *k*PL] across combined [SNR_Pyr_ + *k*PL] clusters. SNR_Pyr_ was significantly different in the low vs. high (Dunn’s multiple comparisons test adjusted *P* < 0.01), and medium vs. high (*P* < 0.001), but not in low vs. medium combined clusters (*P* > 0.99). *k*_PL_ across combined clusters was significantly different in low vs. medium (*P* < 0.01), and low vs. high (*P* < 0.0001), but not between medium vs. high clusters (*P* = 0.20). The normalised mean of the combined [SNR_Pyr_ + *k*PL] metric increased from the low to the high cluster and showed a significant difference in low vs. high comparison (*P* < 0.0001), and medium vs. high (*P* < 0.001), but not in low vs. medium clusters (*P* = 0.06).

Altogether, these results suggest significant inter- and intra-cluster variations in perfusion and metabolism.

### The cluster with the greatest perfusion/metabolism mismatch predicts the highest intratumoral grade

To explore the hypothesis that perfusion/metabolism mismatch may correspond to the most aggressive tumour regions, we assessed the diagnostic performance of each clustering approach in predicting the highest-grade intratumoral region and compared these results to clusters derived from %NG enhancement. The diagnostic performance metrics including sensitivity, specificity, PPV, NPV and AUC of all the clustering approaches for detecting the highest grade within the tumour are shown in Table 2; ROC curves are presented in Fig. 4. Calculations are shown in Suppl. Tables 1 and 2.

**Table 2:** Comparison of the diagnostic performance of the k-means clustering approaches for detection of the highest-grade within an individual tumour. Sensitivity, specificity, positive and negative predictive values (PPV, NPV), and area under the curve (AUC) from ROC curve analysis are presented. The cluster with the highest and most consistent diagnostic performance metrics was the medium combined [SNR_Pyr_ + *k*_PL_] cluster.

| Clustering approach | Sensitivity | Specificity | PPV | NPV | AUC |
| --- | --- | --- | --- | --- | --- |
| **$SNR_{Pyr}$ only** |  |  |  |  |  |
| Low | 40% | 70% | 60% | 52% | 0.58 |
| Medium | 46% | 50% | 50% | 46% | 0.58 |
| High | 23% | 85% | 63% | 50% | 0.54 |
| **$k_{PL}$ only** |  |  |  |  |  |
| Low | 18% | 70% | 40% | 44% | 0.58 |
| Medium | 55% | 80% | 75% | 62% | 0.58 |
| High | 36% | 55% | 47% | 44% | 0.59 |
| **Combined $\text{SNR}_{\text{Pyr}} + k_{\text{PL}}$** |  |  |  |  |  |
| Low | 32% | 65% | 50% | 46% | 0.56 |
| Medium | 64% | 85% | 82% | 68% | 0.67 |
| High | 14% | 55% | 25% | 37% | 0.50 |
| **% Nephrographic enhancement** |  |  |  |  |  |
| Low | 27% | 60% | 43% | 43% | 0.60 |
| Medium | 64% | 65% | 67% | 62% | 0.60 |
| High | 18% | 80% | 50% | 47% | 0.57 |

**Figure 4:**
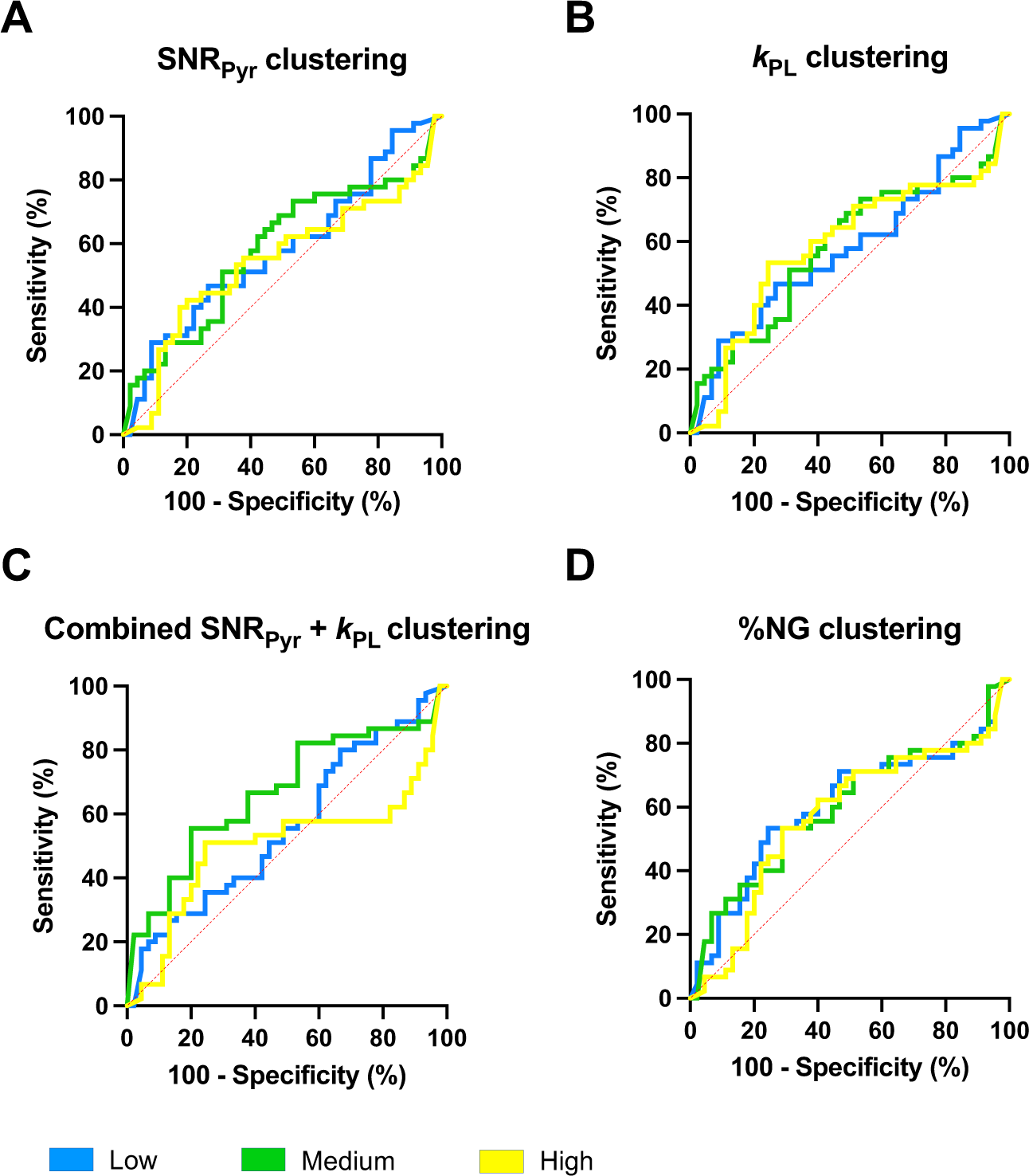
ROC curves for each cluster. (A) SNR_Pyr_ clusters. (B) *k*_PL_ clusters. (C) Combined [SNR_Pyr_ + *k*_PL_] clusters. (D) %NG enhancement clusters. Highest AUC (0.67) was demonstrated by medium combined clustering approach, as seen by green line on ROC curve in (C). Legend: blue line = low clusters; green line = medium clusters; yellow = high clusters.

The medium combined [SNR_Pyr_ + *k*_PL_] cluster exhibited the highest diagnostic performance rates amongst all the clustered parameters with: specificity 85%; sensitivity 64%; PPV 82%; and NPV 68%. Although specificity was comparable to the high SNR_Pyr_ cluster and the sensitivity to the medium %NG cluster, the combined clustering showed the highest consistency, as suggested by ROC curve analysis (AUC 0.67). Therefore, the medium combined [SNR_Pyr_ + *k*PL] cluster, reflecting the low pyruvate delivery and high metabolic conversion (Fig. 3A), demonstrated the best predictor of the highest-grade within the tumour. This prompted further exploration into molecular processes underlying HP ^13^C-MRI clustering.

### The epithelial compartment is the predominant cell type and demonstrates high pyruvate transporter expression suggesting a significant role in HP ^13^C-MRI signal generation

A possible contribution of the tumour microenvironment (TME) and the pyruvate transporter MCT1 to generation of combined clusters was analysed by deconvoluting the bulk RNAseq for cell-type specific signatures, and by quantifying the epithelial and stromal MCT1 staining. Results were compared in binary fashion where the medium combined cluster was compared to others (low + high), as presented on Fig. 5.

**Figure 5:**
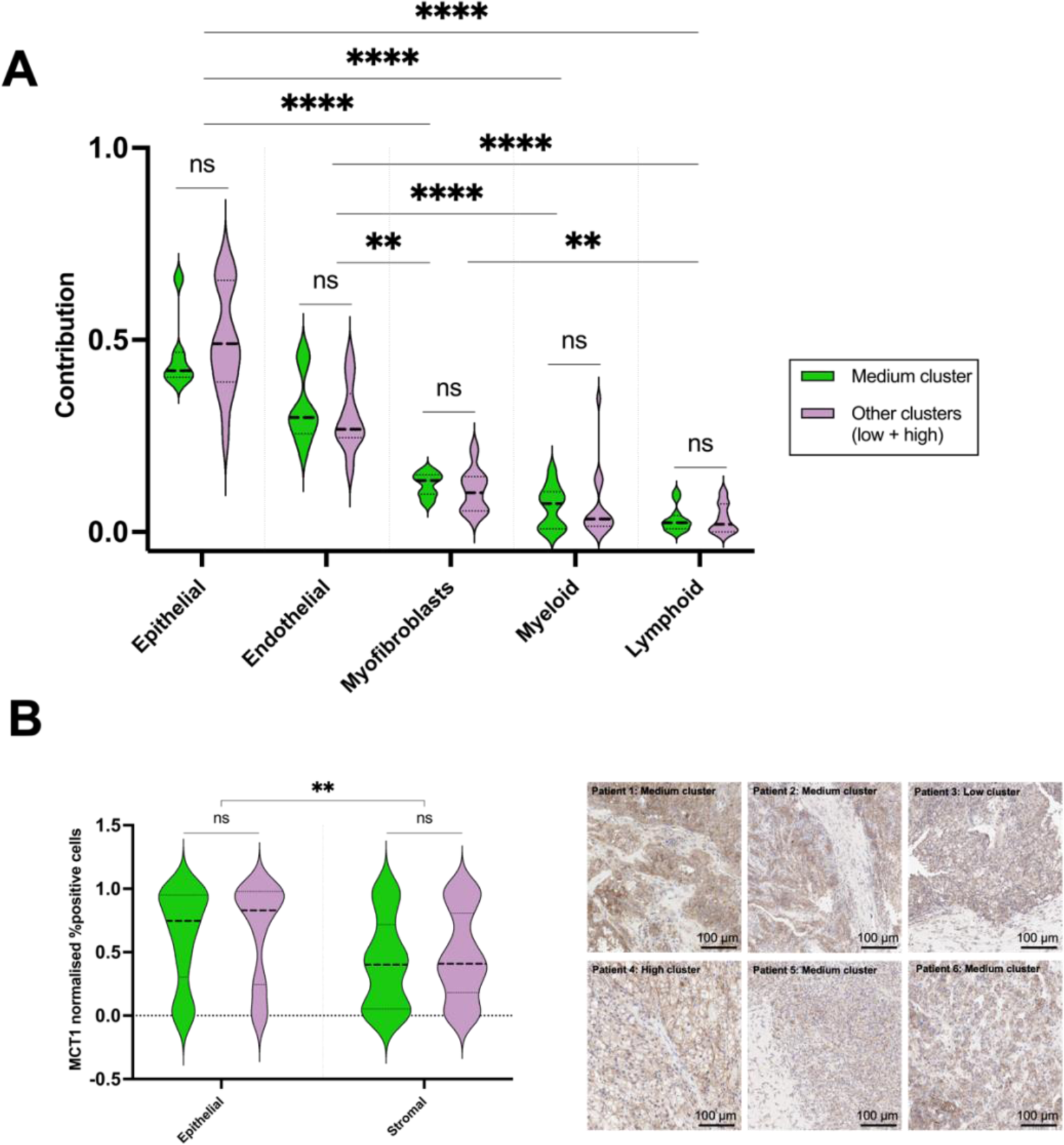
Comparison of tumour microenvironment characteristics within combined clusters. The results were normalised by linear scaling with plots representing: (A) deconvoluted RNAseq identifying cell-type specific signatures, which identified epithelial cells were most abundant compartment, followed by endothelial cells. No significant differences were found comparing medium combined cluster to others. (B) MCT1 expression was significantly higher in epithelial compared to stromal compartment, with representative IHC images from each patient on the right-hand side. No significant differences were found comparing medium combined cluster to others.

Deconvoluted RNAseq-derived cell specific signatures revealed that tumour epithelium contributed the largest relative cellular contribution in the samples, followed by the endothelial component. Both compartments exhibited significantly higher contributions compared to other compartments, as determined by Dunn’s multiple comparisons test (Fig. 5A). However, no difference in cell composition was observed on comparison of the medium combined [SNR_Pyr_ + *k*_PL_] cluster to others.

MCT1 expression was significantly higher in the epithelial compartment compared to the stromal portion (Fig. 5B), but no significant differences were found on intra-cluster comparison. As MCT1 is the main transporter for pyruvate uptake, this is consistent with the epithelial compartment being a major determinant of the HP ^13^C-MRI signal, similar to what has been demonstrated in other tumour types such as prostate cancer^39^. Taken together these results support the hypothesis that the HP ^13^C-MRI signal may be weighted towards metabolism in the epithelial tumour compartment, but between the clusters no TME differences were identified.

### The medium combined cluster revealed the highest metabolic gene expression and aggressive tumour signatures on transcriptomic analysis

To understand the differences in metabolic dysregulation underlying combined [SNR_Pyr_ + *k*_PL_] clusters, GSEA was performed using KEGG MSigDB gene sets. Fig. 7 displays pathways with significance as defined by Benjamini-Hochberg *P*-adjusted values < 0.05 for comparison of medium combined cluster to others (low + high). Suppl. Material S2 contains detailed statistical results.

**Figure 6:**
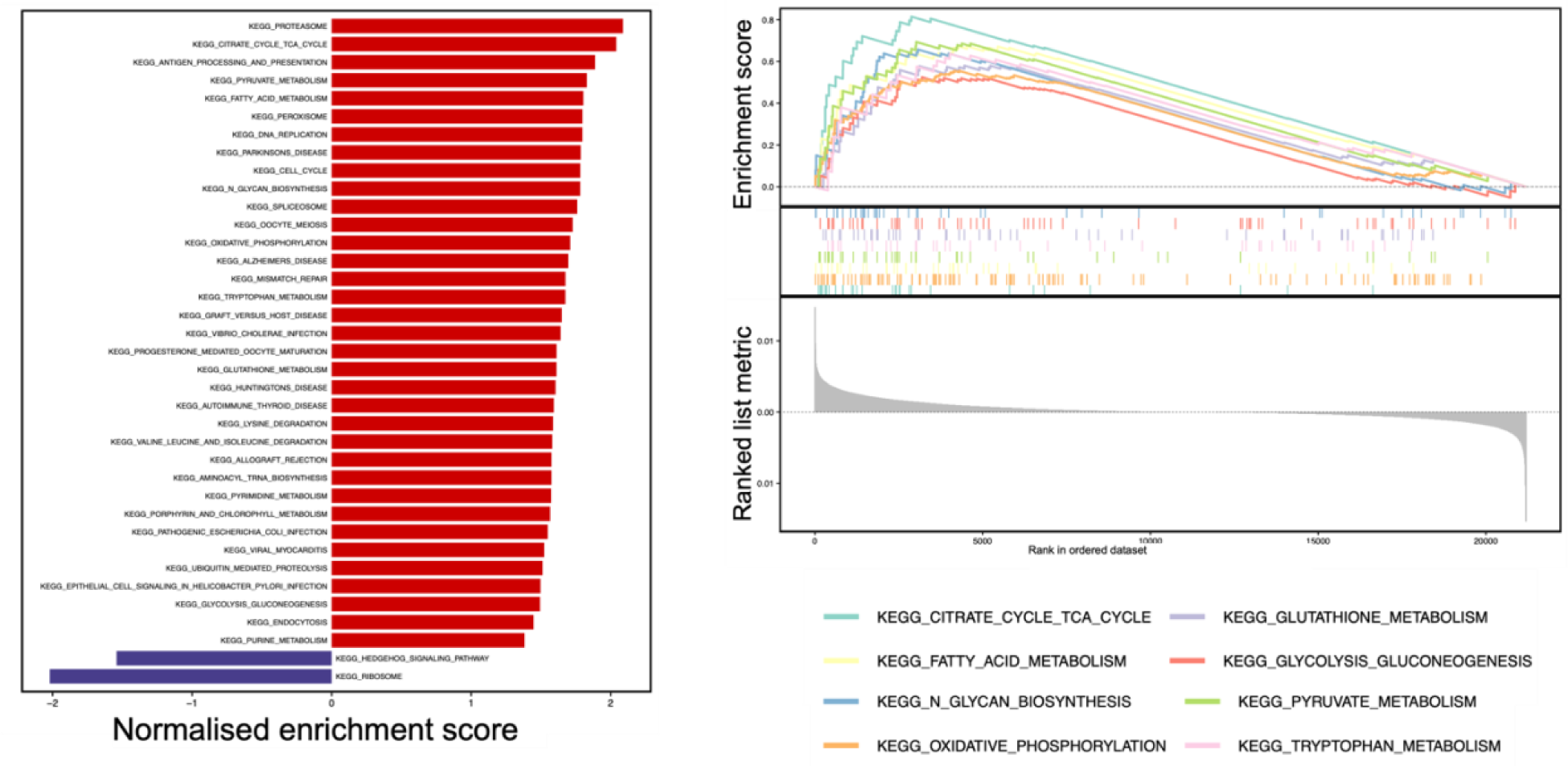
Transcriptomic gene score enrichment analysis for the KEGG MSigDB curated gene set,. (A) Barplots and classic GSEA plots of combined [SNR_Pyr_ + *k*_PL_] clustering, comparing the medium cluster to the others (low + high). Red and blue bars represent up- and down-regulated pathways in medium cluster compared to others, respectively.

**Figure 7:**
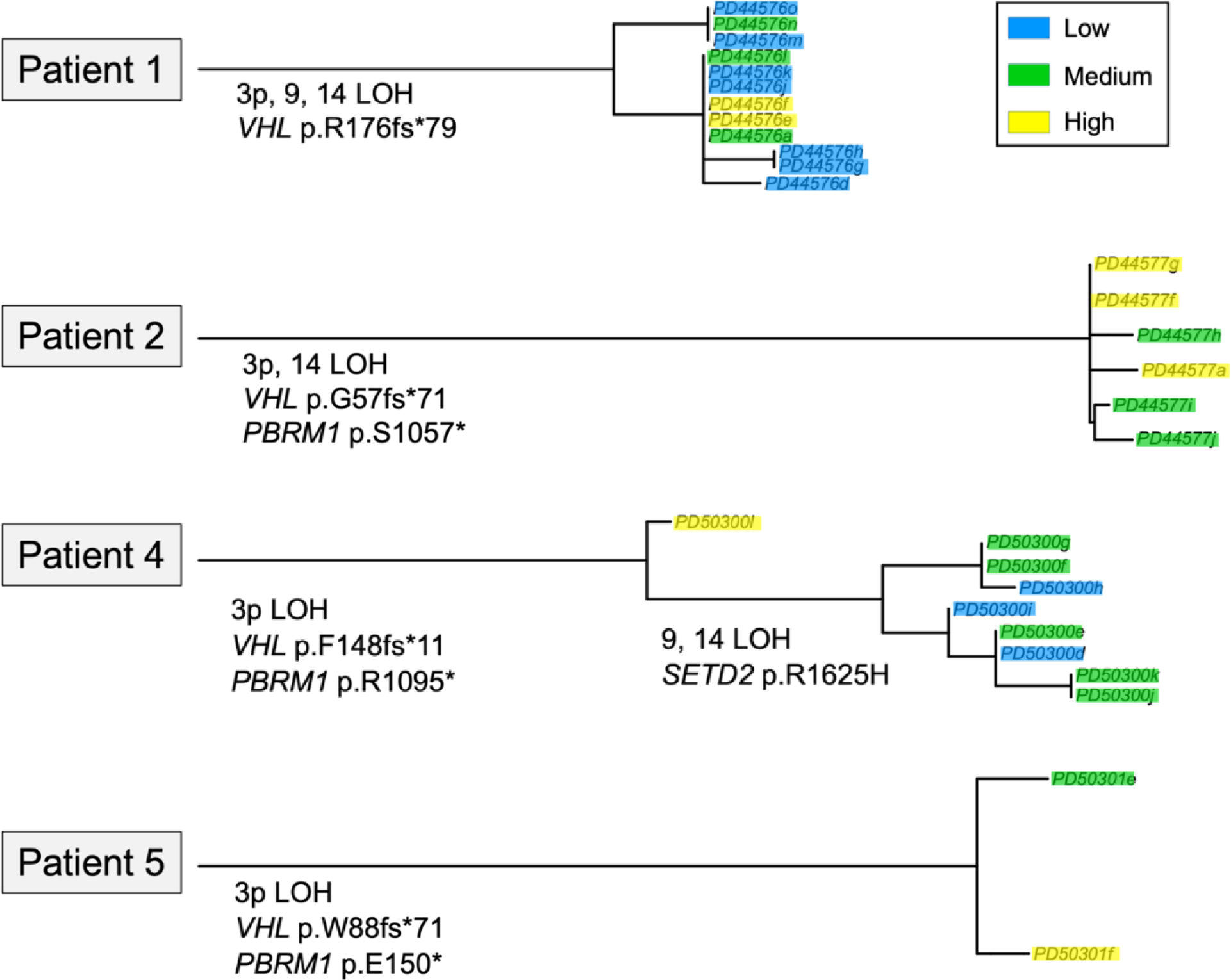
Phylogenies of the genetic alterations in 4 patients. 3p and *VHL* loss were identified as truncal mutations in all patients, with further truncal alterations being 14 loss-of-heterozygosity (LOH) in patients 1 and 4, and *PBRM1* mutation in patients 2, 4 and 5. The only early branching was detected in patient 4, with 9 and 14 LOH, and *SETD2* missense substitution, which was found to underlie all combined medium clusters in this case, with the high cluster harbouring truncal mutation. Legend on top right corner describes colour-coding: blue = low, green = medium, yellow = high cluster.

The medium combined cluster exhibited upregulated metabolic signatures compared to (low + high) combined clusters together. Notably, the energy-yielding metabolic pathways including tricarboxylic acid (TCA) cycle, oxidative phosphorylation (OXPHOS), and pyruvate metabolism ranked among the highest normalised enrichment scores. This was associated with upregulated glutathione and peroxisomal pathways indicating changes in oxidative stress and redox homeostasis. Other significantly enriched metabolic pathways included fatty acid metabolism, N-glycan biosynthesis, glycolysis and gluconeogenesis, and tryptophan metabolism. Upregulation of the proteasome and valine-leucine-isoleucine degradation pathways suggested increased peptide turnover which may be explained by enhanced proliferation, and the requirement for biosynthetic material. In support of this, cell cycle, DNA replication (increased pyrimidine and purine metabolism), and mismatch repair mechanisms were upregulated. The antigen processing and presentation (APP) pathway was also upregulated in the medium cluster compared to others.

### Genetic divergence may partly underlie the aggressive phenotype

Genetic drivers of ccRCC are well-established, with *VHL* loss found in 75% of cases, leading to downstream metabolic phenotype perturbations^40^. To examine the impact of genetic drivers in differentiating the imaging clusters derived from the combined approach, we performed genetic phylogeny analysis, as depicted in Fig. 8. Tissue samples from patients 3 and 6 failed to meet the WES quality control criteria, limiting analysis to the remaining patients.

*VHL* loss was identified as the truncal mutation in all four patients, indicating that it is not a causative factor for intratumoral metabolic heterogeneity. Similarly, 3p loss-of-heterozygosity (LOH) was detected in all patients, and most other genetic alterations were truncal in origin, such as 14q LOH in patient 1 and *PBRM1* alterations in patients 2, 4, and 5. The only exception was the branching of 9, 14 LOH, and the *SETD2* arginine-to-histidine substitution in patient 4, which was notably detected in all samples from the combined [SNR_Pyr_ + *k*_PL_] medium clusters.

## Discussion

Conventional imaging modalities such as CT and MRI are being increasingly used to detect renal masses, but their specificity for determining the presence of ccRCC remains limited^41^. Although biopsy has a high specificity when a diagnostic sample is acquired, non-diagnostic rates, and the potential for undergrading are high (14% and 16% respectively^5^). There is an unmet clinical need for imaging methods to both improve non-invasive diagnosis and to enhance image-guided targeting to the most aggressive intratumoral regions. Metabolic reprogramming is a hallmark of ccRCC and metabolic imaging offers the potential to utilise tumour metabolic phenotypes to improve diagnosis^7^. Here we have employed a HP ^13^C-MRI combined with k-means clustering analysis to investigate whether regional changes in metabolism across the tumour can be used to detect the most aggressive intratumoral regions, with the ultimate aim of guiding biopsy procedures in the future.

To the best of our knowledge, this is the first report of clustering on clinical HP ^13^C-MRI data to define intratumoral metabolic habitats in order to assess their ability to detect highest ccRCC grade within the tumour^42^. In particular, combining the metrics for pyruvate delivery and metabolic conversion to lactate [SNR_Pyr_ + *k*PL] demonstrated the highest diagnostic performance for predicting tumour grade, and it was superior to the clustering approach where each component was used separately or compared to conventional contrast-enhanced MRI. The medium combined cluster showed the highest sensitivity for the detection of aggressive disease and the greatest disparity between metabolism and perfusion. Such perfusion/metabolism mismatch as a feature of tumour aggressiveness is analogous to that reported previously on FDG-PET imaging^36^, but these studies either employed different probes to separately determine blood flow and glucose uptake (e.g. ^15^O-H_2_O and ^18^F-FDG respectively reported in pancreatic^43^, breast^37^ and cervical cancer^38^), or different modalities (e.g. contrast-enhanced CT with FDG-PET in oesophageal cancer^44^). Here we have used a single injection of hyperpolarised ^13^C-pyruvate and HP ^13^C-MRI to extract these parameters within seconds of injection and in the absence of ionising radiation, which is of particular importance in imaging of the kidneys due to renal excretion of ^18^F-FDG which complicates any interpretation^11^. Investigation of perfusion/metabolism mismatch using HP ^13^C-MRI has been reported preclinically using a dual contrast agent approach (^13^C-pyruvate and ^13^C-urea) in a murine prostate cancer model which found a correlation between decreased perfusion and increased metabolism with higher intertumoral grades^45^.

It was previously reported that imaging metrics of pyruvate-to-lactate conversion such as *k*_PL_ and the LAC/PYR ratio positively correlate with higher renal tumour aggressiveness^13,14^. Conventional proton MRI metrics of perfusion have been used to detect higher ccRCC grades e.g. lower arterial spin labelling (ASL)-measured perfusion has been reported in higher ccRCC grades^46^, consistent with lower microvascular density assessed on IHC. However *K*^trans^ as a measure of vascularity on DCE-MRI has shown variability between and within tumours, suggesting inter- and intratumoral heterogeneity^46^. *K*^trans^ is not a pure measure of perfusion but reflects the inflow of the contrast agent from large vessels and its exchange rate into the interstitial space, therefore incorporating a measure of tumour vascular permeability^47^. Here we show that the high cluster of the vascularity parameter, %NG, had the best specificity to predict higher grade region within the tumour, while other metrics varied in their performance. Similarly, Xi *et al.* reported the larger area of high Gd-enhanced k-means cluster within a tumour to be most predictive of higher grade across a cohort of 18 patients with T1b ccRCC^48^; however, these results relied on a single postsurgical grade for the whole tumour rather than examining grade variation across the entire tumour. Udayakumar *et al.* correlated the intratumoral variation in early nephrogenic enhancement with angiogenic and immune transcriptomic signatures of biopsies the same regions^49^ and Yao *et al.* reported two types of microvessels which are differentially expressed between low and high grade ccRCCs, indicating the complexity of ccRCC perfusion^50^. These results indicate that tumour perfusion is an important determinant of aggressiveness but used alone is insufficient to fully and accurately characterise the tumour grade non-invasively. Therefore combining perfusion with other biological processes such as metabolism as we have undertaken here has the potential to improve prediction.

Although intratumoral heterogeneity of pyruvate metabolism in ccRCC has been studied before, this work distinguishes itself not only by evaluating the predictive power of HP ^13^C-MRI-derived metabolic clusters, but also by investigating the underlying biology beyond pyruvate, including cellular compartmentalisation, transcriptomic and genomic signatures. The epithelial cell compartment exhibited the highest expression of pyruvate importer MCT1 and may thus be the driving factor for generation of HP ^13^C-MRI signal as previously suggested^39,51^, but no differences were found between the clusters. Thus, the intratumoural clusters were attributed to variation in metabolic dysregulation as identified on transcriptomic analysis. This is supported by Okegawa *et al.*^10^ who identified differential pyruvate profiles across spatially separated tumour biopsies of eight ccRCC patients, and the high pyruvate cluster was matched by low *LDHA* expression. Through isotope tracing experiments they found higher PDH flux in certain regions, but they have not investigated how this relates to grades or image signal. On the other side, Hakimi *et al.*^52^ and Li *et al.*^53^ identified how ccRCC metabolic clusters are linked to tumour stages and to survival outcomes, but they have only addressed intertumoral rather than intratumoural heterogeneity. Applying HP ^13^C-MRI, Tran *et al.*^54^ reported the highest lactate determined on mass spectrometry (MS) corresponding to the highest ^13^C-lactate signal within a single ccRCC, but have not investigated intratumoural variation of grades or investigated metabolism beyond pyruvate-to-lactate conversion.

Through RNAseq analysis, we identified upregulated metabolic pathways in the medium combined [SNR_Pyr_ + *k*PL] cluster, in particular the notably upregulated TCA cycle pathway. Bezwada *et al.* have previously detected enhanced labelling of TCA intermediates in ccRCC metastases compared to the primary tumour during isotope labelling experiments in patients, suggesting that harnessing of oxidative pathways provides a selective advantage for ccRCC progression^55^. This was further linked to maintenance of redox balance, which contributes to aggressiveness by enabling survival of tumour cells^56^. In our study, the antioxidant-sustaining pathways including glutathione and peroxisome pathways were highly upregulated. Glutathione acts as a scavenger for reactive oxygen species (ROS) to sustain malignant growth^57^, and peroxisomes are also known to regulate lipid droplet formation which is the defining histological feature of ccRCC^58^. Storage of these increased fatty acids is necessary to suppress lipotoxicity and to maintain cell membranes in ccRCC^58^. Glycogen is known to accumulate in these droplets which alters protein glycosylation^59^. Such abnormal glycosylation was shown to promote cell-cell signalling, invasion, and migration^60^, and we have shown increased N-glycan biosynthesis in the most aggressive combined cluster in this study. The tryptophan metabolic pathway was the most prominently upregulated among amino acid metabolic pathways, which is well known in ccRCC for immunosuppressive effects via the kynurenine pathway^61^. In the medium combined [SNR_Pyr_ + *k*PL] cluster, the cell proliferation pathways were upregulated, which is tightly linked to higher cancer aggressiveness and used in clinical routine histological grading of ccRCC that is based on nucleolar prominence^62^.

This study is the first to compare gene aberrations to metabolic phenotypes detected on HP ^13^C-MRI in ccRCC. Intratumoral genomic clonality of ccRCC has been extensively studied^63,64^, but it remains unclear how this translates to a variation in metabolic phenotypes with evidence suggesting that it is only partly related to genetic alterations^65^. We identified truncal *VHL* loss in all the patients and although this is a major driver of the difference in metabolism between tumour and normal tissue, it is unlikely to contribute to the intratumoral metabolic heterogeneity detected on HP ^13^C-MRI. This finding was consistent with Okegawa *et al.*, who reported intratumoral variation of pyruvate-to-lactate conversion on mass spectrometry that was independent of *VHL* gene status^10^. While most other genetic alterations were truncal, the single exception of genetic branching was the LOH of chromosomes 9 and 14, and *SETD2* mutation in patient 4, which were all found within the combined medium cluster. Indeed, LOH of chromosomes 9 and 14, as well as elevated genomic intratumoural heterogeneity, have been previously associated with more adverse outcomes^66–68^, which supports our suggestion that the medium combined cluster represents the most aggressive intratumoral region. In addition, *SETD2* loss has been previously found to promote ccRCC expansion through replication stress and defective DNA damage repair^69^, as well as a switch of ccRCC metabolism towards OXPHOS^70^, in line with our finding of upregulated OXPHOS pathway in the medium combined cluster. Although this was the first attempt to link intratumoral genomic variations with metabolic clusters on imaging, no direct causation was observed due to the limited number of patients. Future studies will need to further elucidate the extent to which additional factors, such as epigenetics and noncanonical metabolic flux, regulate the metabolic phenotype.

Our research faced unavoidable technical challenges: HP ^13^C-MRI images have inherently low resolution (voxel size: 17 × 17 × 30 mm^3^), which may dilute the border between heterogeneous regions especially in the superior-inferior direction. This was the reason for allocating biopsies from two different axial levels of tissue sampling onto the same HP ^13^C-image of study patient 5. Therefore, only 2D-analysis of HP ^13^C-MRI was possible. Also, biopsy sampling was performed only on 2D slices of nephrectomised kidneys since whole-tumour tissue analyses are technically challenging and impossible in a clinical setting due to the need for diagnostic samples. Still, multiregional sampling with the 3D-printed tumour mould enabled good correspondence between imaging and pathology^22^, and by clustering of ^13^C-data we mitigated the effects of any possible misregistration, therefore providing a larger confidence space for potential future biopsy targeting. In summary, we present a novel clustering method of HP ^13^C-MRI data that predicts the most aggressive intratumoural ccRCC grades with high specificity, outperforming individual parameters and the clinical standard. This work supports the potential of metabolic imaging to guide biopsy to the most aggressive tumour regions, thereby reducing sampling error and RCC undergrading, improving risk stratification and clinical management strategies. Additionally, the analysis of underlying molecular processes sets the ground for future research into intratumoral metabolic heterogeneity of ccRCC with potential for development of novel treatments.

## Acknowledgements

This study was funded by Cancer Research UK (C19212/A27150, C19212/A29082, C19212/A16628). This research was supported by the Mark Foundation for Cancer Research (RG95043), Cancer Research UK Cambridge Centre (C9685/A25177 and CTRQQR-2021\100012), the NIHR Cambridge Biomedical Research Centre (BRC-1215-20014 and NIHR203312). The views expressed are those of the authors and not necessarily those of the NIHR or the Department of Health and Social Care. The authors have additional funding from the National Cancer Imaging Translational Accelerator (NCITA; C42780/A27066), the Cambridge Experimental Cancer Medicine Centre, the Mark Foundation Institute for Integrated Cancer Medicine (MFICM), and the Canadian Institute For Advanced Research (CIFAR).

