## Supplementary for "K-means clustering of hyperpolarised ^13^C-MRI identifies intratumoural perfusion/metabolism mismatch in renal cell carcinoma as best predictor of highest grade"

| Biopsy | Grade | Highest grade? 1/0 | $SNR_{Pyr}$ | $k_{PL}$ | [$SNR_{Pyr}$+$k_{PL}$] | %NG |
|---|---|---|---|---|---|---|
| MRE_001_T1 | 4 | 1 | M | M | M | L |
| MRE_001_T2 | 3 | 0 | L | M | L | L |
| MRE_001_T3 | 3 | 0 | H | H | H | H |
| MRE_001_T4 | 3 | 0 | M | H | H | H |
| MRE_001_T5 | 3 | 0 | L | H | L | L |
| MRE_001_T6 | 3 | 0 | L | M | L | L |
| MRE_001_T7 | 2 | 0 | M | H | H | H |
| MRE_001_T8 | 3 | 0 | M | M | L | M |
| MRE_001_T9 | 2 | 0 | M | M | L | M |
| MRE_001_T10 | 3 | 0 | M | L | M | M |
| MRE_001_T11 | 3 | 0 | L | L | L | M |
| MRE_001_T12 | 4 | 1 | M | M | M | M |
| MRE_001_T13 | 4 | 1 | L | M | L | H |
| MRE_001_T14 | 4 | 1 | M | H | M | M |
| MRE_006_T1 | 2 | 0 | M | L | H | H |
| MRE_006_T2 | 1 | 0 | M | L | H | M |
| MRE_006_T3 | 2 | 0 | L | L | H | L |
| MRE_006_T4 | 2 | 0 | H | H | M | M |
| MRE_006_T5 | 3 | 1 | M | M | M | M |
| MRE_006_T6 | 3 | 1 | M | M | M | L |
| MRE_010_T1 | 1 | 0 | H | H | H | M |
| MRE_010_T2 | 1 | 0 | L | L | L | L |
| MRE_010_T3 | 1 | 0 | M | H | H | L |
| MRE_010_T4 | 2 | 1 | M | H | M | M |
| MRE_010_T5 | 2 | 1 | L | L | L | M |
| MRE_012_T1 | 4 | 1 | H | M | L | M |
| MRE_012_T2 | 4 | 1 | M | H | M | L |
| MRE_012_T3 | 4 | 1 | L | H | M | M |
| MRE_012_T4 | 4 | 1 | L | M | M | H |
| MRE_012_T5 | 4 | 1 | H | L | L | M |
| MRE_012_T6 | 4 | 1 | H | M | L | L |
| MRE_012_T7 | 4 | 1 | M | H | M | M |
| MRE_012_T8 | 4 | 1 | L | M | M | H |
| MRE_012_T9 | 2 | 0 | M | H | H | L |
| MRE_014_T1 | 2 | 1 | L | H | M | M |
| MRE_014_T2 | 2 | 1 | L | H | M | M |
| MRE_014_T3 | 2 | 1 | M | M | H | L |
| MRE_014_T4 | 2 | 1 | L | L | L | M |
| MRE_014_T5 | 1 | 0 | M | H | M | L |
| MRE_014_T7 | 2 | 1 | H | M | H | M |
| MRE_016_T1 | 2 | 1 | L | L | M | H |
| MRE_016_T2 | 2 | 1 | L | M | H | M |
| MRE_016_T3 | 2 | 1 | M | M | L | L |
| MRE_016_T4 | 2 | 1 | H | H | H | H |

**Supplementary Table 1: Allocation of biopsies to corresponding clusters.**

| | | | | | | | | | | |
|---|---|---|---|---|---|---|---|---|---|---|
| | YES Highest grade | YES Highest grade | | | | | YES Highest grade | NO Highest grade | | |
| SNRPyr L+ | 9 | 6 | PPV | 0.6 | | kPL L+ | 4 | 6 | PPV | 0.4 |
| SNRPyr L- | 13 | 14 | NPV | 0.51851852 | | kPL L- | 18 | 14 | NPV | 0.4375 |
| | Sensitivity | Specificity | | | | | Sensitivity | Specificity | | |
| | 0.40909091 | 0.7 | | | | | 0.18181818 | 0.7 | | |
| | YES Highest grade | NO Highest grade | | | | | YES Highest grade | NO Highest grade | | |
| SNRPyr M+ | 10 | 10 | PPV | 0.5 | | kPL M+ | 12 | 4 | PPV | 0.75 |
| SNRPyr M- | 12 | 10 | NPV | 0.45454545 | | kPL M- | 10 | 16 | NPV | 0.61538462 |
| | Sensitivity | Specificity | | | | | Sensitivity | Specificity | | |
| | 0.45454545 | 0.5 | | | | | 0.54545455 | 0.8 | | |
| | YES Highest grade | NO Highest grade | | | | | YES Highest grade | NO Highest grade | | |
| SNRPyr H+ | 5 | 3 | PPV | 0.625 | | kPL H+ | 8 | 9 | PPV | 0.47058824 |
| SNRPyr H- | 17 | 17 | NPV | 0.5 | | kPL H- | 14 | 11 | NPV | 0.44 |
| | Sensitivity | Specificity | | | | | Sensitivity | Specificity | | |
| | 0.22727273 | 0.85 | | | | | 0.36363636 | 0.55 | | |
| | YES Highest grade | NO Highest grade | | | | | YES Highest grade | NO Highest grade | | |
| Combined3 L+ | 7 | 7 | PPV | 0.5 | | %NG L+ | 6 | 8 | PPV | 0.42857143 |
| Combined3 L- | 15 | 13 | NPV | 0.46428571 | | %NG L- | 16 | 12 | NPV | 0.42857143 |
| | Sensitivity | Specificity | | | | | Sensitivity | Specificity | | |
| | 0.31818182 | 0.65 | | | | | 0.27272727 | 0.6 | | |
| | YES Highest grade | NO Highest grade | | | | | YES Highest grade | NO Highest grade | | |
| Combined3 M+ | 14 | 3 | PPV | 0.82352941 | | %NG M+ | 14 | 7 | PPV | 0.66666667 |
| Combined3 M- | 8 | 17 | NPV | 0.68 | | %NG M- | 8 | 13 | NPV | 0.61904762 |
| | Sensitivity | Specificity | | | | | Sensitivity | Specificity | | |
| | 0.63636364 | 0.85 | | | | | 0.63636364 | 0.65 | | |
| | YES Highest grade | NO Highest grade | | | | | YES Highest grade | NO Highest grade | | |
| Combined3 H+ | 3 | 9 | PPV | 0.25 | | %NG H+ | 4 | 4 | PPV | 0.5 |
| Combined3 H- | 19 | 11 | NPV | 0.36666667 | | %NG H- | 18 | 16 | NPV | 0.47058824 |
| | Sensitivity | Specificity | | | | | Sensitivity | Specificity | | |
| | 0.13636364 | 0.55 | | | | | 0.18181818 | 0.8 | | |

**Supplementary Table 2: Diagnostic performance calculations.**
